# TRANSITION FROM CLINICAL PRACTITIONER TO ACADEMIC EDUCATOR: THE EXPERIENCES OF RADIOGRAPHY LECTURERS IN NIGERIA

**DOI:** 10.1101/2023.11.14.23298531

**Authors:** Michael Promise Ogolodom, H.U. Chiegwu, Awajimijan Nathaniel Mbaba, Abdul Fatai K. Bakre, Elizabeth O. Balogun, Annmaris Chimebere Obasi, Uchenna Norochukwunso Ezechukwu, Emeka E. Ezugwu, Egop Brownson Egop

## Abstract

**Background** Clinical educators are essential in radiography education programmes. Transitioning from clinical practicing radiographers to academic educators is a rewarding journey. This study was designed to examine the experience of Radiography Lecturers in the Nigerian universities that converted from radiography practitioners to academic educators. **Materials and methods:** A Google form questionnaire-based study design was conducted among 35 radiography lecturers in Nigeria. Only lecturers in Nigerian Universities who had worked as radiography clinical practitioners before transited to academics and consented to participate in this study were included. The questionnaire captured responses on socio-demographic variables, challenges, barriers, successes of transition from practitioners to academic educators. **Results:** The majority 17(48.6%) of the respondents earned income of above #251,000.00 as a practicing radiographers. Of the 35 respondents, the majority 12(34.3%) each respectively earned #101,000.00 −#150,000.00 and #151,000.00 - #200,000.00 as an academic radiographers. Majority 17(48.6%) of the respondents agreed that the main challenge they are encountering as an academic educator is rigorous research activities. Family issues affected smooth transition process as 23(65.7%) of the respondents agreed to that. Majority 18(51.4%) perceived mentorship as the key factor responsible for smooth transition from practice to academic. There was statistically significant relationship between gender and challenges encountered by the responders (χ^2^ = 28.194, p = 0.00). **Conclusion:** The respondents experienced different challenges as they transited from clinical practice to academic. Mentorship is the key factor that militated against the smooth transition process. There was statistically significant relationship between gender and challenges encountered by the responders.

## Introduction

Radiological science education and training is of paramount clinical importance given the prominence of medical imaging utilization in effective clinical practice. The digital revolution has fueled advancements in radiography, which has positively impacted radiology education and training (Majumder *et al.,* 2021). Globally, standard radiography training at the undergraduate degree level combines classroom instruction, laboratory practice of clinical skills, and clinical placement. These essential training elements are created to provide students with a range of learning opportunities, necessary information, skills, and competencies to practice safely and successfully after graduation. Medical professionals who want to share their clinical and radiographic skills with others just starting out in the field often choose to be teaching as a rich and satisfying career option. Teaching offers the chance to impact student progress and mold the upcoming generation of technologists and radiographers (Azer, 2005). A gratifying path is taking the step from working as a radiographer to becoming an academic.

Transitioning from clinical practicing radiographer to an academician is a rewarding journey (Thompson and Taylor, 2020). It requires a new level of professional development and commitment to ensure radiography students receive a well-prepared clinical learning experience (Chamuyonga *et al.,* 2020). It is important for novice radiography academics to acquire skills and competencies that will provide radiography students with quality and effective learning environments (Lawal *et al.,* 2022). Clinical education is the mainstay for skill development and it has been demonstrated that combining practical and theoretical education leads to a significantly better outcome in the field of teaching. This integrated approach to knowledge and practice in education enables the trainee to work more competently and to be ready to take on responsibility in his or her future career (Deputy of Organization Development and Human Resource Management [DOHRM], 2003). Clinical educators are essential in radiography education programmes. Clinical teachers, according to McLeod *et al*. (2009), have practical "how to do it" teaching knowledge, but few are familiar with the fundamental ideas, theories, and concepts underlying the teaching and learning process or the why behind pedagogical behaviors. This study sought to examine the experience of Radiography lecturers in the Nigerian University setting that converted from radiography clinical practice to academic educators.

## Materials and Methods

This study conducted among radiography lecturers in Nigeria Universities was a cross-sectional questionnaire-based survey. Only lecturers in Nigeria Universities who had worked as radiography clinical practitioner before transited to academics and consented to participate in this study were included. Radiography lecturers in Nigeria Universities who has not practiced before joining the academics were excluded from the study. The purpose of this study was duly explained to the respondents and their consent was sought and obtained using written informed consent form. Their privacy and confidentiality were assured and they were at liberty to withdraw from the study at any time without any harm. An ethical approval **(FHST/REC/023/00101)** was obtained from the Human Research and Ethical Committee of the Faculty of Health Sciences and Technology, Nnamdi Azikiwe University, Nnewi Campus, Anambra, State, Nigeria.

The sample size for the study was determined using the statistical formula for unknown population used by previous study Charan and Biswas(2013).

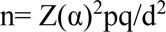

Where;

n=expected sample size

Zα = significant level usually set at 95% confidence level, Zα is 1.96 (two sided)

p= portion of the population with similar attributes under study = 50% (0.5)

d= margin of error tolerated or absolute error = 16. 6% (0.166) q= 1-p=1-0.5= 0.5

Therefore; n= 1.96^2^ ×0.5x0.5 / 0.166^2^

n= (0.9604) / (0.027566)

n = 34.8≈ 35

A random sampling technique was used to select the 35 participants. The consent of the participants were duly sought and obtained using written informed consent.

A Self-constructed questionnaire was employed as the instrument for data collection. The questionnaire was structured into five sections; A, B, C, D, and E based on the specific objectives of the study: Section A consists of demographic variables of participants detailing 7-questions. section B entail the factors that militate against the transition processes, sections C detail the challenges, section D detail the factors responsible for the smooth transition from practitioner to academic educator. Section E details the successes/benefits of being an academic educator.

### Validity and reliability Test

A pilot study was carried out with 10 questionnaires distributed among radiography lecturers before the study commences. The Cronbach alpha reliability test was calculated and the questionnaire had an acceptable internal consistency (Cronbach’s alpha = 0.91). The validity of the questionnaire was determined using the Index of term Objective Congruence (IOC) method used by previous authors (Turner and Carlson, 2013; Mbaba *et al*.,2021; Ogolodom *et al*., 2020). This was done by calculating the index of item-objective congruence (IOC). According to the index parameters, an IOC score higher than 0.6 was assumed to show adequate content validity, and all the scores obtained in this study for all the items of the questionnaire after IOC analysis was higher than 0.6

The questionnaire was produced in electronic and hardcopy versions. The electronic version was issued using WhatsApp groups for Radiography Lecturers Association of Nigeria (RLAN), and Nnamdi Azikiwe University Radiography Lecturers and the respondent’s feedback was retrieved electronically. The hardcopy version was distributed by the researchers using one-on-one method and retrieved immediately after completion. This study was conducted from December 2022 to March 2023.

The data collected were analyzed using the Statistical Package for Social Sciences software (SPSS) version 21.0(IBM Corp, Armonk, NY, 2012). Data obtained from the study were analyzed using descriptive statistics (frequencies, percentages, tables and charts) and inferential statistics (Chi-square). Values were considered significant at *p<0.05*.

## Results

### Socio-demographic characteristics of the respondents

Out of 30 respondents, males were 30(85.7%) when compared with their female counterparts 5(14.3%). Most of the respondents 18(51.4%) had 3-10 years practice experience. The majority 18(51.4%) had 3-10 years academic experience and the least 5(14.3%) had 11-20 years of academic experience. The majority 17(48.6%) of the respondents earned income of above #251,000.00 as a practicing radiographers and the least 6(17.1%) respondents earned income of the range #101,000.00 - #150,000.00.Of the 35 respondents, the majority 12(34.3%) each respectively earned #101,000.00 −#150,000.00 and #151,000.00 - #200,000.00 as an academic radiographers(Table 1). Greater number 11(31.4%) of the respondents were in the age group of 46-50years and the least were in the age ranges of 30-35 years and 36-40years, which is 6(17.1%) each respectively(Figure 1). Large proportion 23(65.7%) of the respondents had Ph.D in radiography and the remainder had MSc, which is 12(34.3%)(Figure 2).

**Figure 1:**
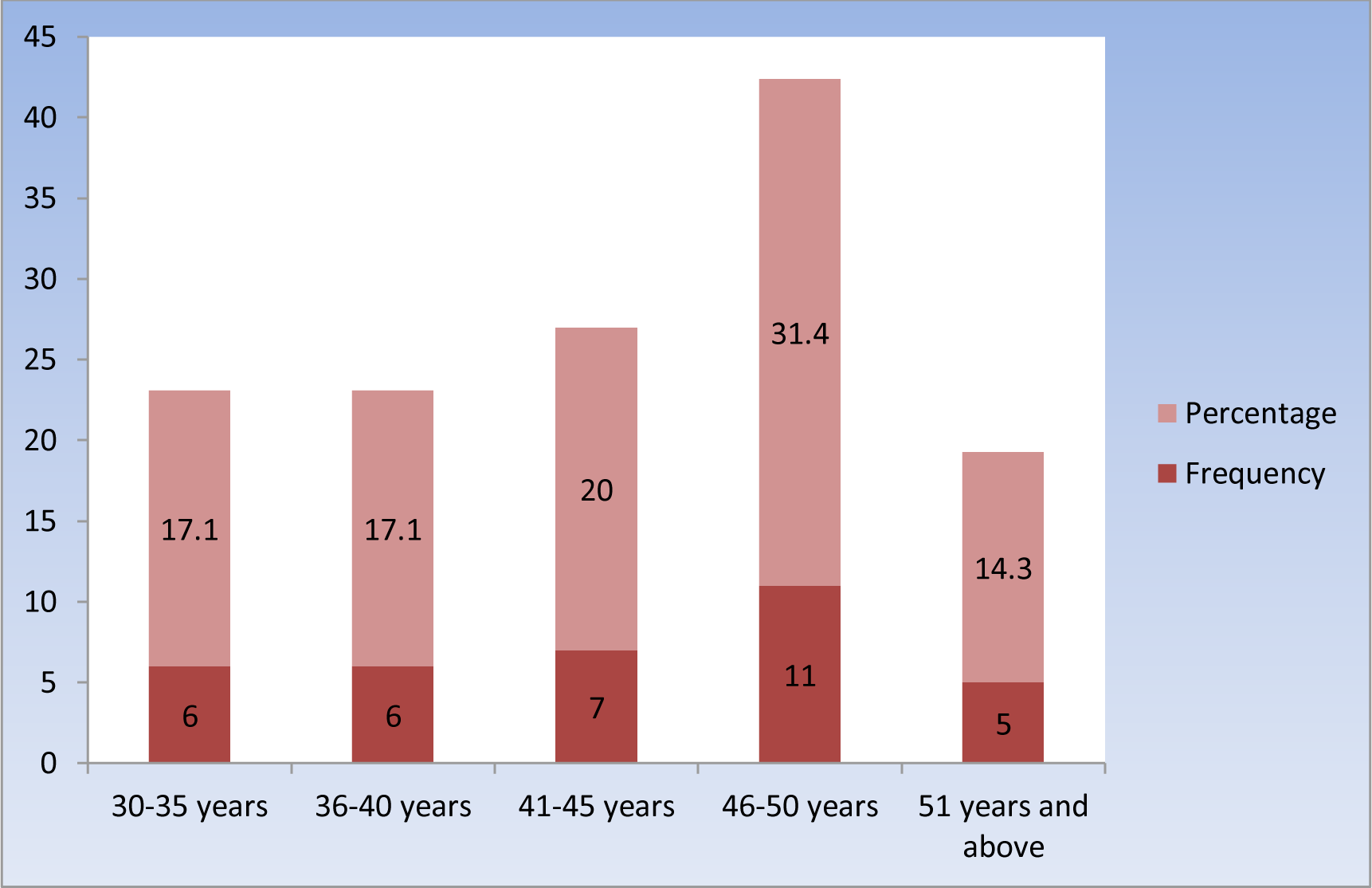
Bar chart showing frequency and percentage of age group.

**Figure 2:**
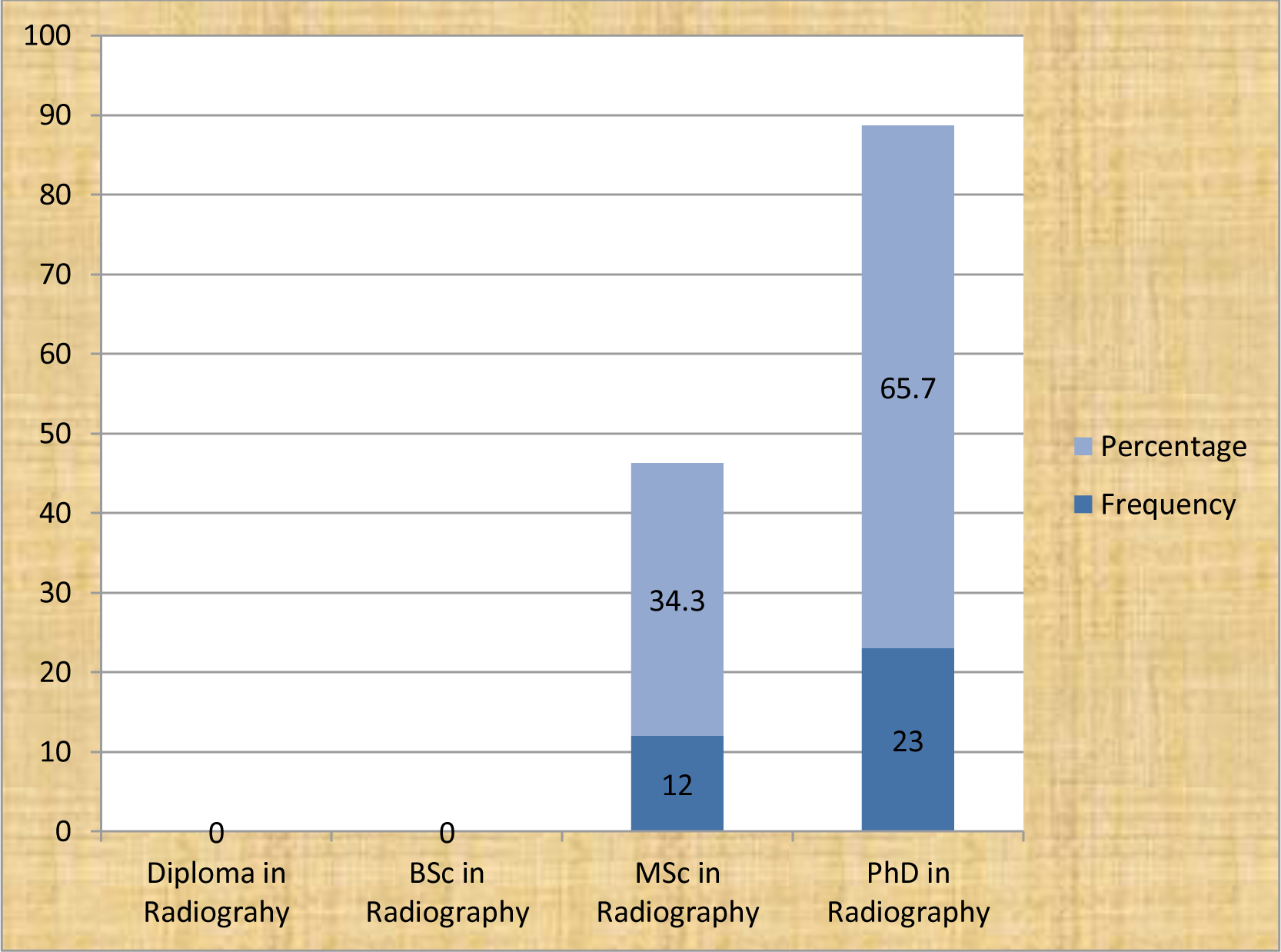
Bar chart showing frequency distribution of educational level.

**Table 1:** Showing Socio-demographic Characteristics of participants.

| Variables | Frequency(F) | Percentage (%) |
| --- | --- | --- |
| **Gender** |  |  |
| Male | 30 | 85.7 |
| Female | 5 | 14.3 |
| Total | 35 | 100.0 |
| **Years of Practicing Experience** |  |  |
| < 2 years | - | - |
| 3-10 years | 18 | 51.4 |
| 11-20 years | 17 | 48.6 |
| >20 years | - | - |
| Total | 35 | 100.0 |
| **Years of Academic Experience** |  |  |
| < 2 years | - |  |
| 3-10 years | 18 | 51.4 |
| 11-20 years | 5 | 14.3 |
| >20 years | 12 | 34.3 |
| Total | 35 | 100.0 |
| **Income earned in Practicing Radiographer** |  |  |
| Less than #100,000 | - | - |
| #101,000-#150,000 | 6 | 17.1 |
| #151,000-#200,000 | 12 | 34.3 |
| #201-#250,000 | - | - |
| Above #251,000 | 17 | 48.6 |
| Total | 35 | 100.0 |
| **Income earned as an Academic Radiographer** |  |  |
| Less than #100,000 | - |  |
| #101,000-#150,000 | 12 | 34.3 |
| #151,000-#200,000 | 12 | 34.3 |
| #201-#250,000 | - | - |
| Above #251,000 | 11 | 31.4 |
| Total | 35 | 100.0 |

### The factors that militated against smooth transition from clinical practicing radiographer to an academic educator

Most 23(65.7%) of the respondents agreed that family issues affects their smooth transition from clinical practice to academics. Greater number 23(65.7%) of respondents agreed that lack of proper education can affect the smooth transition process. Out of 35 respondents, 24(68.6%) agreed that the educational qualification level has effect on the transition process from clinical practice to academics (Table 2).

**Table 2:**
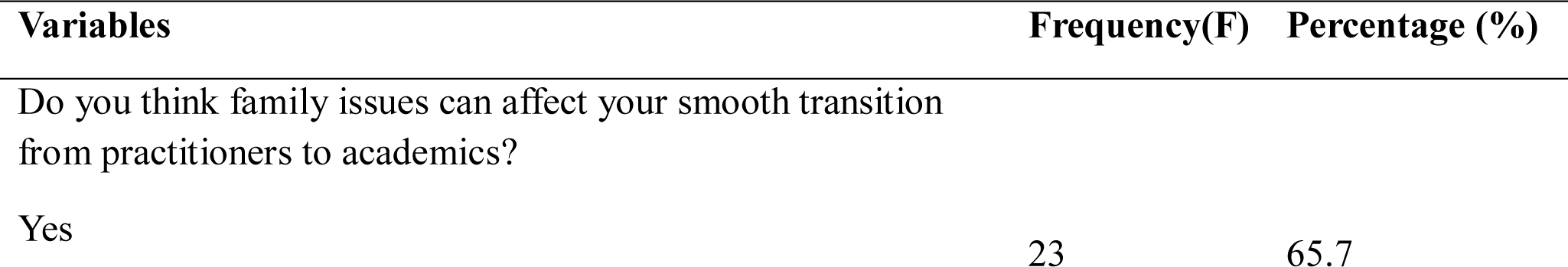
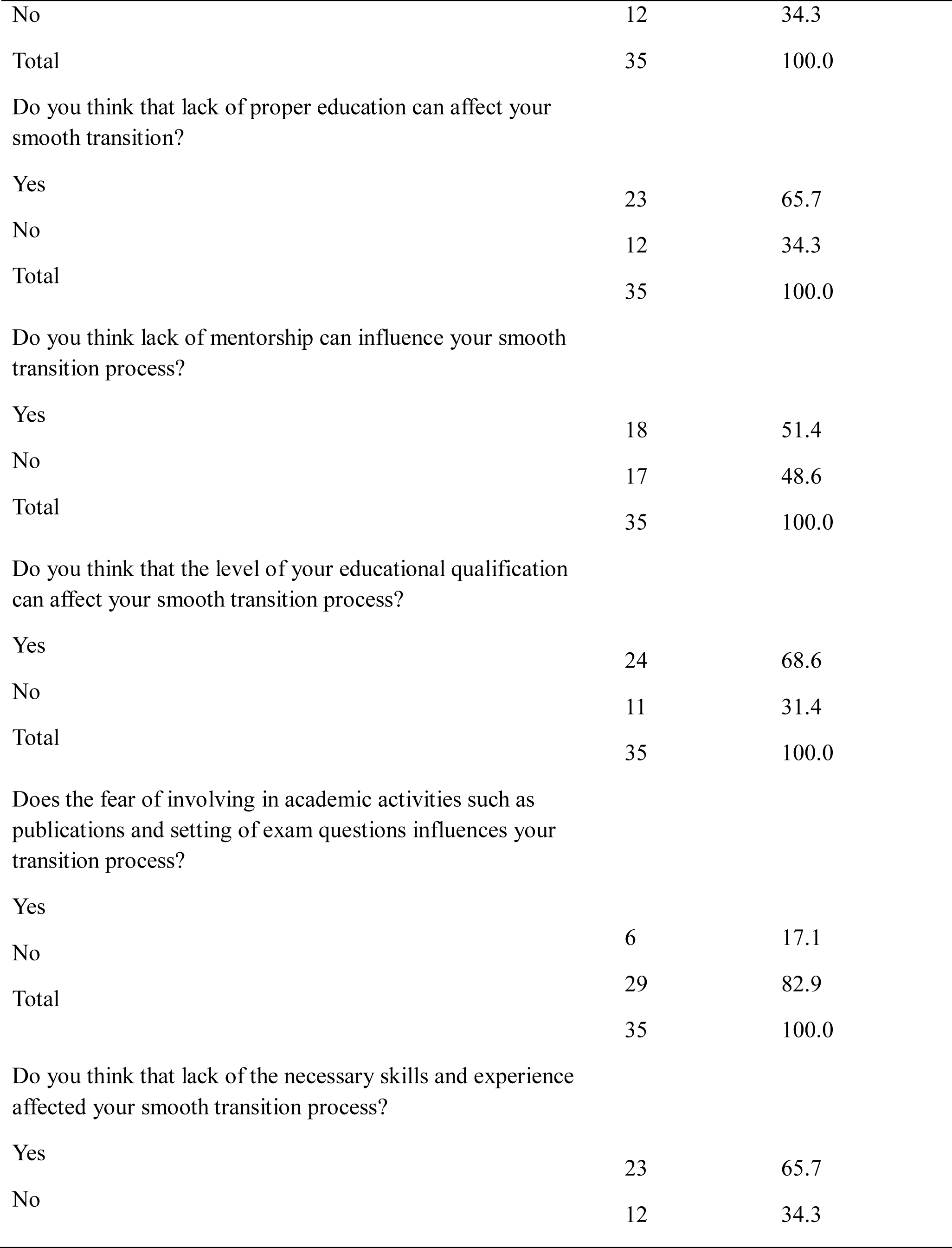
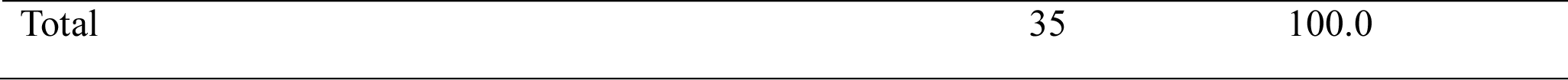
Factors that militated against smooth transition from practitioner to an academic educator.

### The challenges encountered by Radiography lecturers that transited from clinical practitioner to academic educators

The majority 17(48.6%) of the respondents strongly agreed that the main challenge encountered as an academic educator was rigorous research activities and the least 6(17.1%) disagreed. Out of 35 respondents, 18 (51.4%) agreed that most conferences and workshops were mostly self-sponsored while 17(48.6%) strongly agreed (Table 3a). Greater proportion 23(65.7%) of the respondents strongly agreed that academic educators are mostly involved in the preparation of lecture materials, marking of exams scripts and evaluating of students via assessment test, which are tedious component of academic activities. The majority 18(51.4%) of the respondents agreed that poorly funded research and inadequate laboratory equipment were also some of their challenges as academic educator (Table 3b).

**Table 3.**
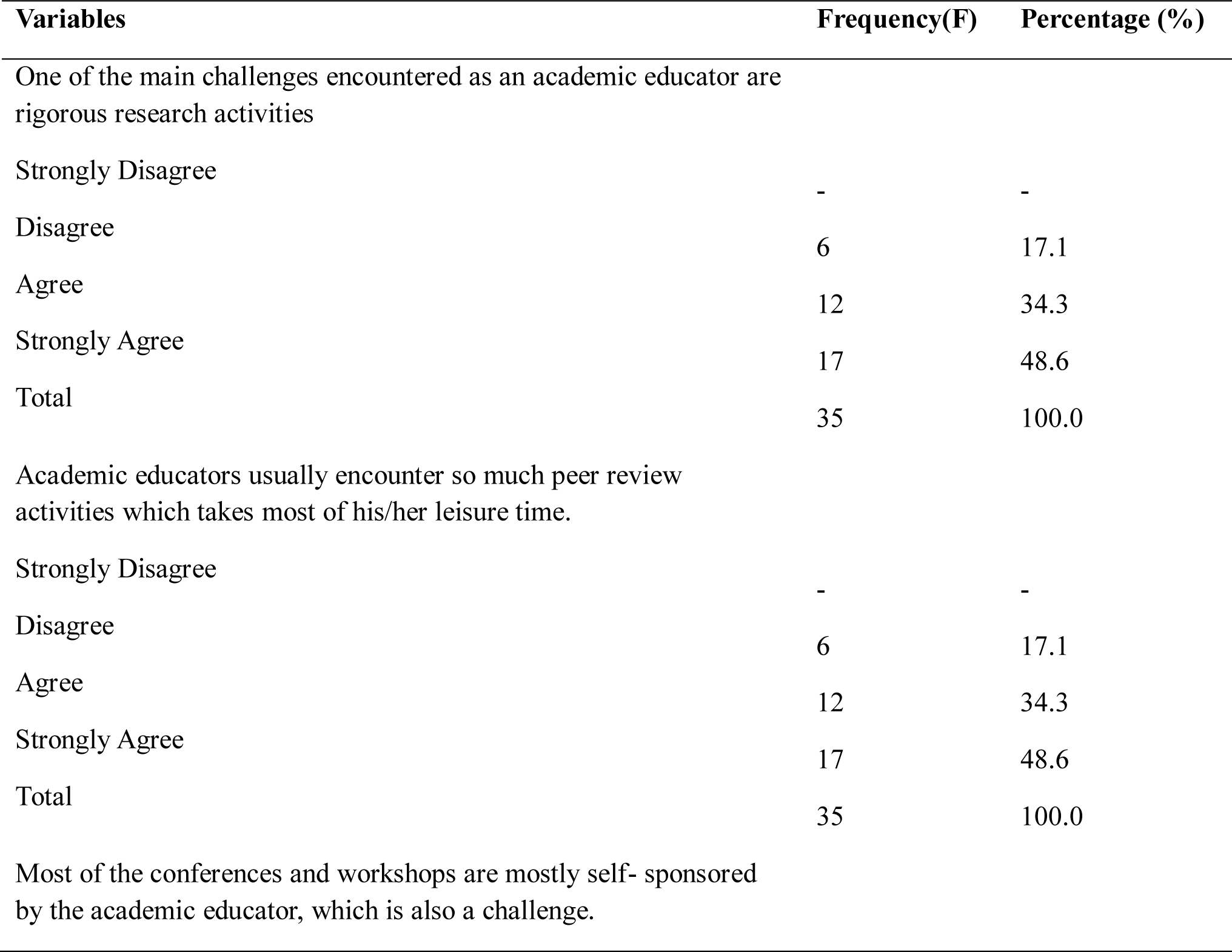
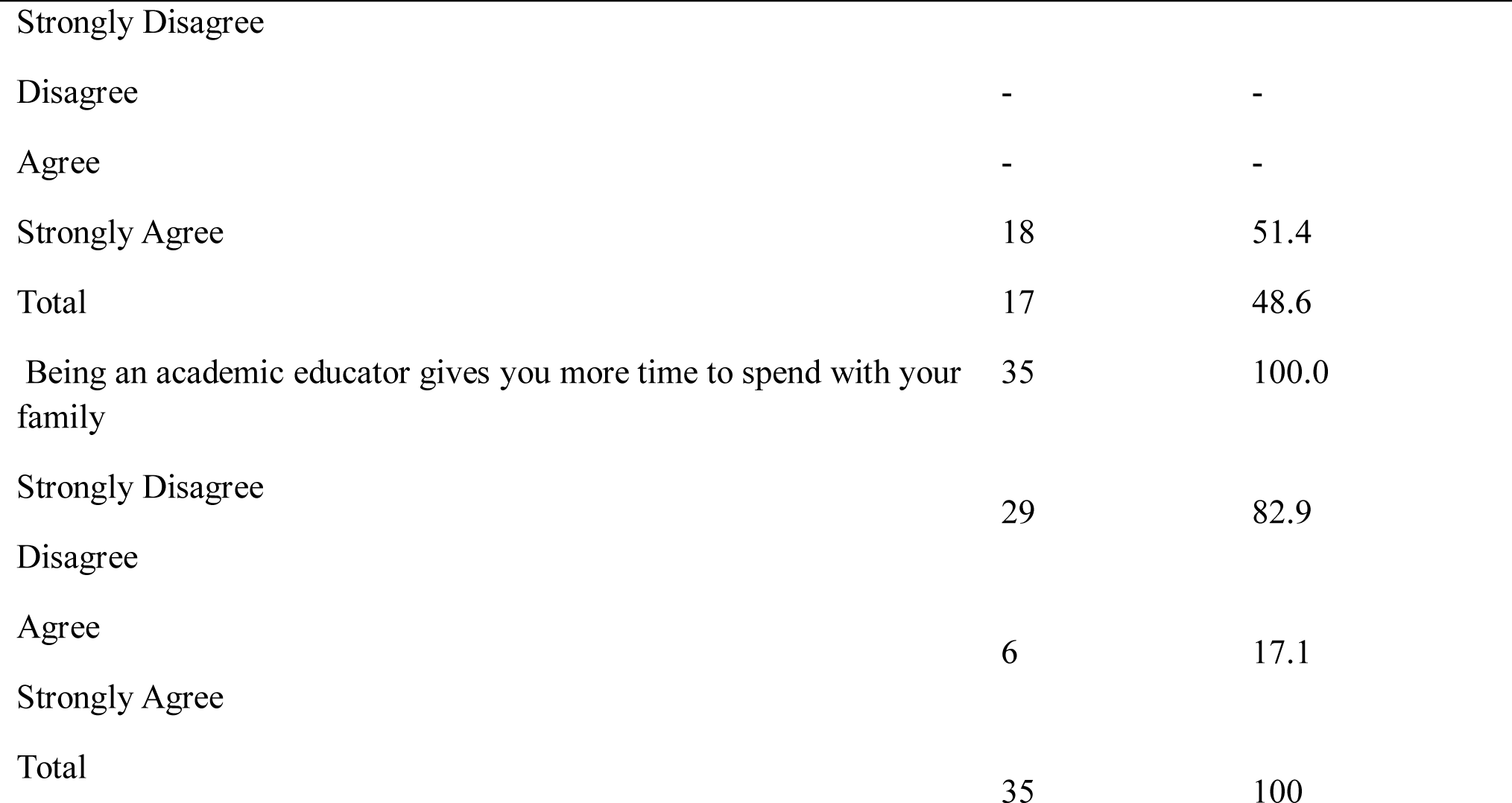
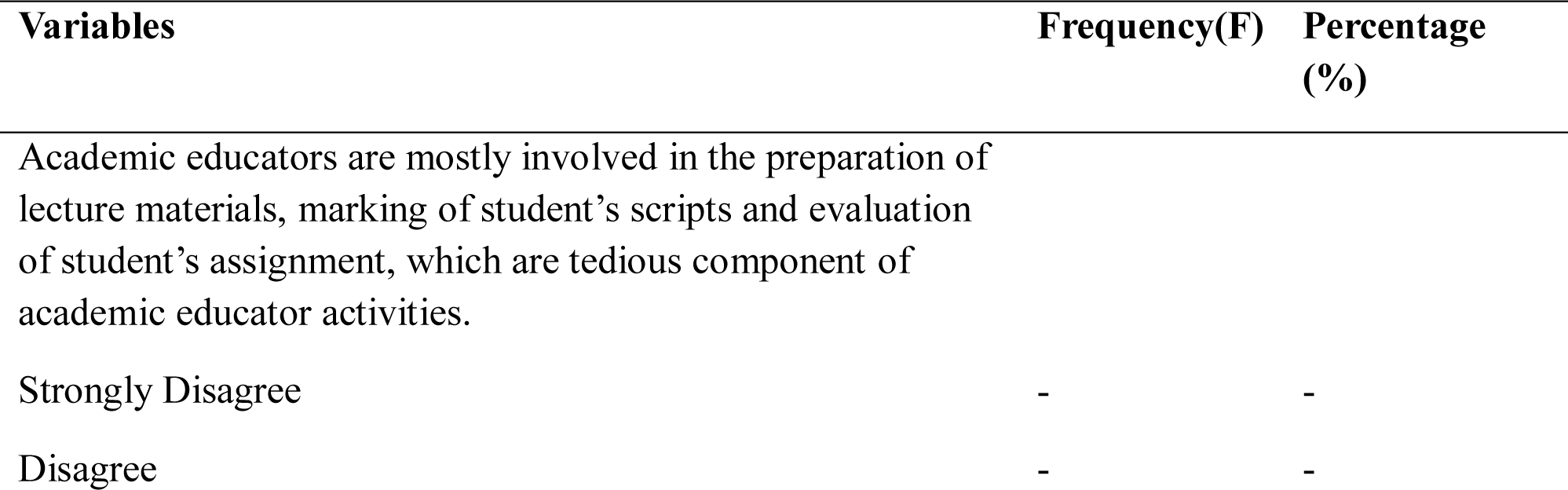
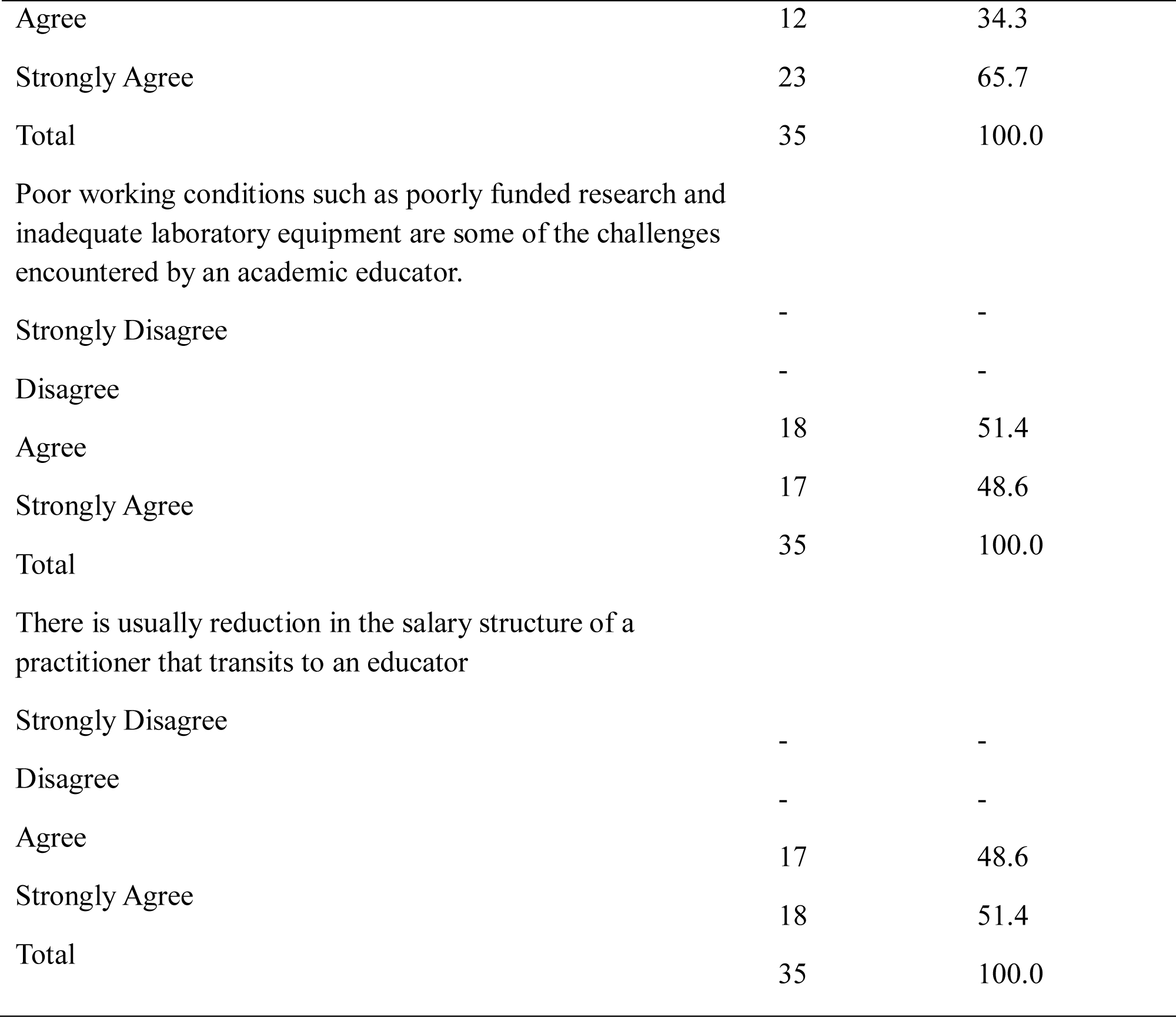
a: Challenges encountered by radiography lecturers that transited as a practitioner to an academic educator b: Challenges encountered by radiography lecturers that transited as a practitioner to an academic educator

### The factors responsible for the smooth transition from clinical practicing radiographer to an academic educator

Out of 35 respondents, 18(51.4%) agreed that mentorship was responsible for their smooth transition from clinical practice to academics. Large number 18(51.4%) of the respondents disagreed that closeness to schools offering Radiography programme could affect their smooth transition process (Table 4).

**Table 4:**
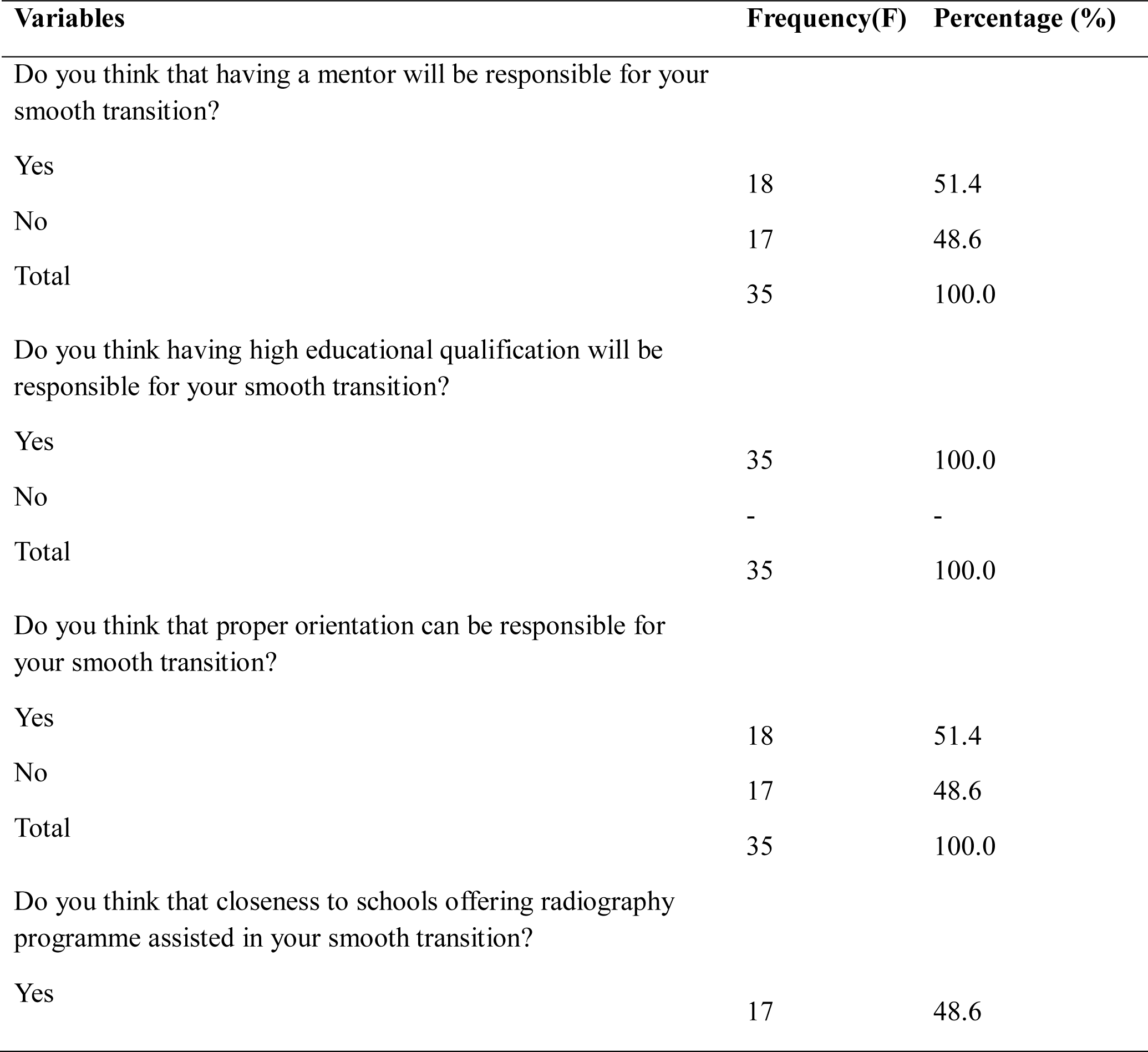
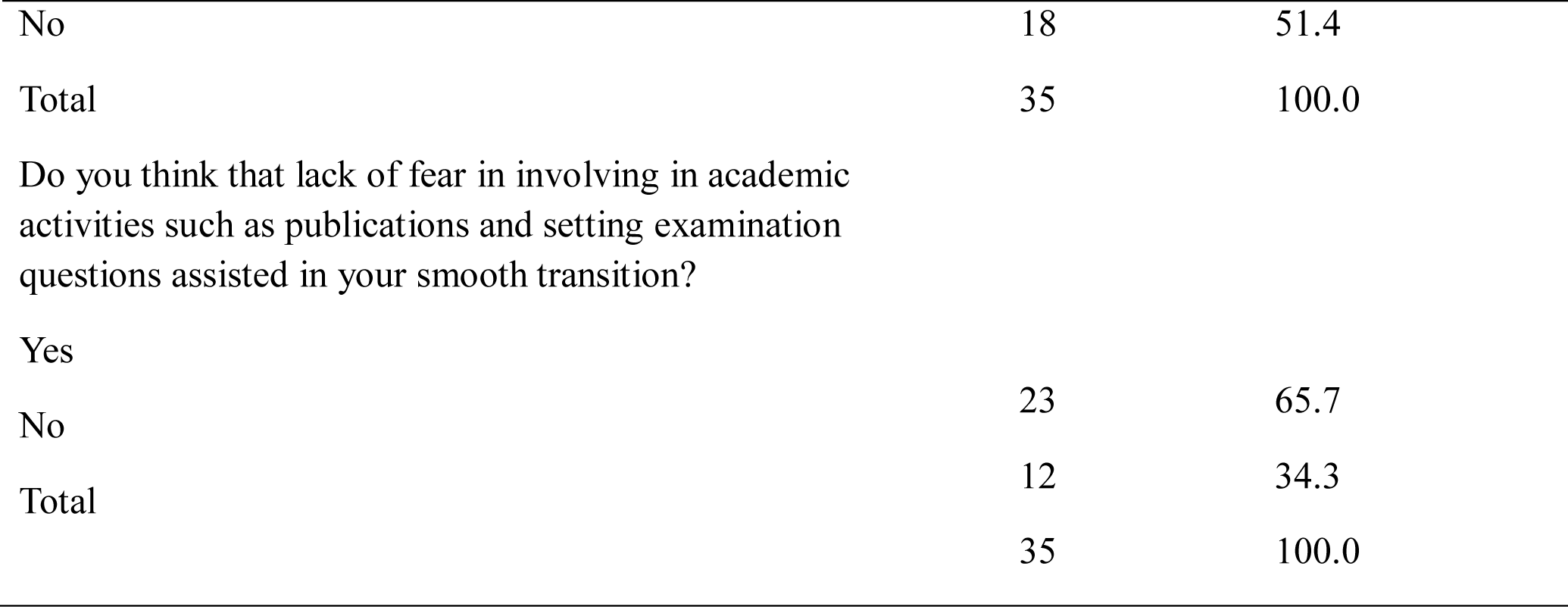
Frequency and percentage distribution of factors responsible for smooth transition.

### The successes/benefits of being an academic educator

Of the 35 respondents, 29(82.9%) perceived themselves being successful as academic educator. All, 35(100%) the respondents, perceived their transition from clinical practice to academics as a blessing to Radiography profession. The majority 29(82.9%) of the respondents, perceived their societal status to have increased since they became academic educators. Out of 35 respondents, 30(85.7%) agreed that they are impacting on more lives now than in clinical practice (Table 5).

**Table 5:** Success and benefits of being an academic educator.

| **Variables** | **Frequency(F)** | **Percentage (%)** |
| --- | --- | --- |
| Do you think you are a successful academic educator? |  |  |
| Yes | 29 | 82.9 |
| No | 6 | 17.1 |
| Total | 35 | 100.0 |
| Are there known benefits associated with being academic educators? |  |  |
| Yes | 29 | 82.9 |
| No | 6 | 17.1 |
| Total | 35 | 100.0 |
| Do you think that transiting from practice to academics would be of benefit to radiography profession? |  |  |
| Yes | 35 | 100.0 |
| No | - | - |
| Total | 35 | 100.0 |
| Do you think that being an academic educator have increased your status in your community or society at large? |  |  |
| Yes | 29 | 82.9 |
| No | 6 | 17.1 |
| Total | 35 | 100.0 |
| As an academic educator, do you think you are impacting on more lives than in practice? |  |  |
| Yes | 30 | 85.7 |
| No | 5 | 14.3 |
| Total | 35 | 100.0 |

### The relationships between the participants’ gender and the challenges encountered during the transition process from clinical practice to academics

There was a statistically significant relationship between the respondents’ gender and the challenges encountered during the transition process (χ^2^ = 28.194, df= 4, p = 0.000)(Table 6).

**Table 6:**
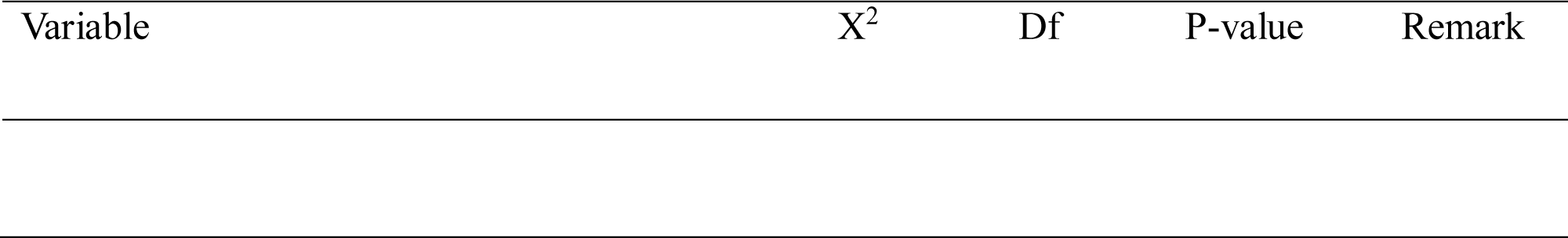
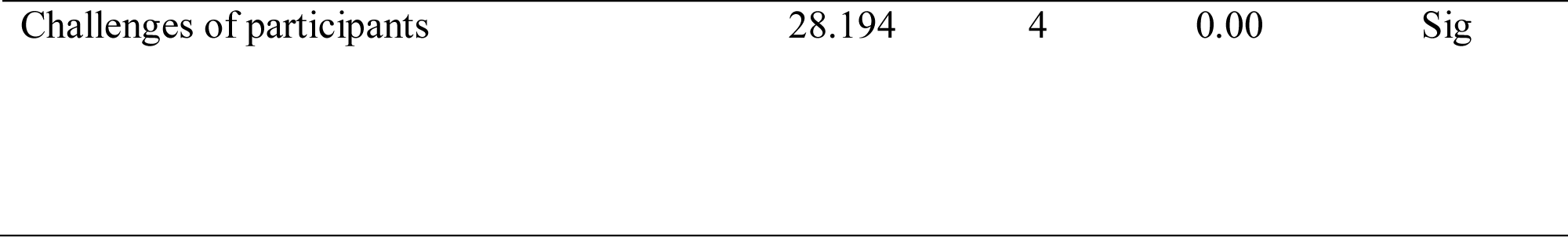
Chi-Square table showing relationship between the participant’s gender and challenges encountered during the transition process.

| Variable | $X^2$ | Df | P-value | Remark |
| --- | --- | --- | --- | --- |
| Challenges of participants | 28.194 | 4 | 0.00 | Sig |

## Discussion

Despite pre-registration education and service organization modifications, reality shock sets in practice, which remains a problem for healthcare professionals. The shift from clinical practice to academic educator is a critical stage for diagnostic radiographers as they begin independent practice in a demanding setting and continue to establish their professional identity (Burton *et al*., 2015; Morley, 2009).

In this index study, males were the majority when compared with their female counterparts. This contracted the greater number of females than males reported by Maduka *et al*. (2021), Ma *et al* (2022) among radiographers, and also by Barnes (2015) among clinical nurses who transited to academics.

The results on the factors that militated against smooth transition from clinical practice to academics revealed that most of the respondents agreed that family issues affected their smooth transition. Greater number of respondents agreed that lack of proper knowledge can affect the smooth transition process. Over 60% of the respondents agreed that educational qualification level has effect on the transition process from clinical practice to academics which agreed with the result obtained by Knapp *et al* (2022). This implies that some clinical radiographers may have had the interest of moving to academics, but due to some of the aforementioned factors. This finding may present the reality as most radiographers settled for clinical practice with their first degree, which is B.Sc due to the high financial benefits associated with clinical practice.

From the responses from respondents, there are numerous challenges encountered by clinical radiography as they transited to academic educators. These were collaborated by many other resaech findings and include but not limited to rigorous research activities (Thompson and Taylor, 2020), most conferences and workshops were self-sponsored (Laari *et al*., 2021), involvement in academic activities, poor working conditions, that being an academic educator does not give more time to spend with their family and reduction of salary earned by a clinical radiographer who transits to be an academic educator. This agrees with the finding by Laari *et al*. (2021) who reported that workshops and conferences help build their confidences and eases to academic educator despite been self-sponsor, which is in line with the study.

This shows that radiography academic educators are faced with varied challenges, which may differ according to institutions and individuals. These findings are also in agreement with the studies conducted by Kyei *et al*. (2015), which reported that the preparation for clinical training by academic educator was significant.

With regards to the factors that are responsible for the smooth transition process, the majorities of the respondents agreed that mentorship, higher educational qualifications, proper orientation and courageously engaging in academic activities were some key factors responsible for their smooth transition process. Also, good number of them disagreed that closeness to schools offering radiography programmes did not affect their transition process. These findings imply that having a mentor, higher educational qualifications, proper orientation and lack of fear to partake in academic activities are the essential ingredients a clinical radiographers needs to smoothly transit to academics. The study findings have agreement with the reports of McDermid et al. (2016), Heale *et al* (2009), Dempsey (2007) and Gardner (2007) revealing that good mentorship by senior colleagues would ease smooth transition process.

The result of the study on the success and benefits of being an academic educator indicated that the majority of the respondents were successful academic educators while very insignificant number perceived them not successful. This finding could be based on the individual concept of success, as what one called success may not be the same for another person. Also, all the respondents perceived their transition from clinical practice to academics as beneficial to the radiography profession. This could be seen from the fact that currently in Nigeria, there are so many schools offering Radiography programmes but there is inadequate number of academic radiographers, which could be attributed to the fact that most radiographers preferred the clinical practice (where they earn more salary) to academics. Therefore, those that transited from clinical practice to academics are really a blessing to the radiography profession, since they could help to train more manpower needed in both clinical setting and academics.

In addition, most of the respondents perceived their status in the society to increase due to their position as academic educators. Most of the respondents, have agreed that by being academic educators they have impacted on more lives than in clinical practice. This finding was similar to the statements documented by Thompson and Taylor (2020) and Ryan and Deci (2000) in their research work, which revealed that as academic educator they were exposed to different avenues employed in impacting the lives of the students.

The result on the relationships between the respondent’s gender and the challenges encountered due to their transition from clinical practice to an academic educator revealed that there was statistically significant relationship between the respondent’s gender and the challenges encountered due to their transition from clinical practice to an academic educator. This finding implies that degree of challenges male respondents encountered differs from that of their female counterparts. This may be so because most of the female academics were married women and so they would have so many things distracting their attentions, thereby making working as academic educators more tedious to them than being in clinical practice.

## Conclusion

Transiting from clinical radiographers to academic radiographers in Nigeria presents a lot of challenges due to a number of factors ranging from loss in salary, lack of the proper knowledge of what are involved, lack of mentorship, the task of self-sponsored workshops and conferences, family responsibilities, educational qualification and so on and so forth. However, there are the perceived gains of impacting positively on more lives, increased social status and contributing to enhance the radiography profession

## Author’s contributions

MPO and ACO were the main researchers, drafted the manuscript and captured the data. HUC, MPO, ACO, ANM, AKB, EOB, UNE, EEE and EEB analysed the results HUC, MPO, ACO, ANM, AKB, EOB, UNE,EEE and EBE gave recommendations on the review of the literature and were critical readers of the work. All authors have read and approved the final manuscript.

## Data Availability

All data produced in the present work are contained in the manuscript

## Acknowledgement

Authors acknowledge all the respondents who participated in this study

